# Association of white matter hyperintensities, white matter microstructural changes, hippocampal and amygdala volumes with neuropathology in a community cohort using 7T postmortem in situ MRI

**DOI:** 10.64898/2026.06.30.26356455

**Authors:** Jr-Jiun Liou, Maria da Graca Morais Martin, Roberta Diehl Rodriguez, Lea T. Grinberg, Tales Santini, Tamer S. Ibrahim, Maria Concepción García Otaduy

**Affiliations:** University of Pittsburgh, Pittsburgh, PA 15213, USA; University of Sao Paulo, Sao Paulo, SP 05508-220, Brazil; Mayo Clinic, Jacksonville, FL 32224, USA

**Keywords:** Fazekas score, diffusion, mean diffusivity, medial temporal atrophy score, neuritic plaque

## Abstract

**INTRODUCTION:** We integrate visual and quantitative metrics in white matter and medial temporal lobe to examine relationships with neuropathology in a community cohort.

**METHODS:** Postmortem in situ MRI (T1, T2, DWI) was performed in 25 human brains, followed by visual ratings (Fazekas, MTA, ERICA, Koedam). Neuropathology included ADNC, LATE-NC, hippocampal sclerosis, PART, ARTAG, Lewy pathology, and CAA.

**RESULTS:** Increased Fazekas score was linked to aging, lower education, hypertension, higher basilar artery wall thickness, and greater Braak NFT stage. WMH volume also correlated with lacunes, Thal phase, and CERAD score that was not observed using Fazekas. Hippocampal volumes were lower in elderly, less educated people and were associated with higher atrophy scores and higher Braak NFT stage. Higher amygdala volume was only associated with higher CERAD score.

**DISCUSSION:** Quantitative MRI may detect neuropathologic associations more sensitively than visual ratings. Tau pathology is a key predictor of WMH burden and hippocampal atrophy.

## Introduction

Brain autopsies remain the gold standard for accurately diagnosing neurodegenerative and cerebrovascular diseases. However, certain neuroimaging findings, such as focal edema and white matter hyperintensities (WMHs), are difficult to identify on gross tissue sections without radiological guidance. Postmortem magnetic resonance imaging (MRI) at 7 tesla (7T) offers an opportunity to: (i) scan and evaluate the whole brain, while most of the neuropathological studies are based on tissue samples of limited regions; (ii) capture end-stage WMHs more accurately than antemortem MRI, which is often acquired up to a decade before autopsy; and (iii) provide significantly higher image contrast and resolution compared to clinical MRI at 3 tesla.

Why do WMHs matter? Increased WMHs are associated with aging and are classically linked to cerebral small vessel disease and lower cognitive scores ^1^. They are also associated with greater Alzheimer’s disease (AD) neuropathology, including increased cortical phosphorylated tau immunostaining ^2, 3^. Interestingly, increased total WMH volume is associated with increased cerebrospinal fluid total tau in those younger than 65 years, but not in those older than 65 years^4^. Beyond global WMH burden, regional patterns are important. Frontal lobe WMHs can help differentiate frontotemporal lobar degeneration with TDP-43 pathology from AD pathology ^5^. Deep frontal WMH is associated with hypertension and diabetes whereas smoking has no significant association ^6^. Higher WMHs in juxtacortical and deep frontal regions are associated with two major cerebrovascular pathologies - cerebral amyloid angiopathy (CAA) and arteriolosclerosis, respectively ^7–10^. Higher WMH volumes in the parietal lobe are associated with increased risk of AD ^11–14^ as well as axonal loss and demyelination ^7–9, 15–19^. One large radiological-pathological correlation study found that greater WMH burden is associated with higher arteriolosclerosis severity and presence of gross infarcts in those with no cognitive impairment ^20^. Interestingly, a recent study shows WMH volume is not associated with the severity of arteriolosclerosis in CAA-confirmed cases ^21^. These contradicting observations are likely due to the differences between cohort selection.

Beyond AD, WMH burden also correlates with atrophy in the fronto-insular and medial temporal regions in primary age-related tauopathy (PART), particularly in the fronto-insular region and Fazekas score can differentiate between PART and AD ^22^. Other PART studies also show that TDP-43 pathology is associated with greater volume loss in the hippocampus and amygdala ^23, 24^ and that increasing Braak neurofibrillary tangle stage is linked to atrophy in the anterior and medial temporal regions ^25, 26^. In cases with limbic-predominant age-related TDP-43 encephalopathy neuropathologic change, amygdala and hippocampal atrophy are also observed and that the medial temporal atrophy (MTA) score has the same predictive capabilities as postmortem hippocampal volume ^27–33^.

Clinically, WMH burden is commonly assessed using the Fazekas score, which is visually rated by neuroradiologists. Another modality, diffusion-weighted imaging (DWI), is not only sensitive to restricted diffusion caused by cytotoxic edema or tumor growth, but also sensitive to increased diffusion caused by neurodegenerative processes. One of the measurable outcomes from DWI, mean diffusivity (MD), has been shown to differentiate between healthy controls and patients with multiple sclerosis ^26^. Hippocampal and amygdala atrophy are clinically evaluated visually using the MTA score by neuroradiologists. We therefore wonder whether quantitative volumetric and diffusivity assessments can provide more granular information than the Fazekas and MTA score. In this study, we combined multimodal measures for visual and quantitative assessment to investigate the association of WMH, white matter microstructural changes, hippocampal and amygdala volumes with neuropathology in a community cohort.

## Methods

### Study population and clinical diagnosis

Our study was approved by the Institutional Review Board (IRB) at the University of Sao Paulo in Sao Paulo, Brazil (IRB approval CAAE: 95570518.2.0000.0065). The inclusion criteria were older adults aged 50 years or older. The consent was obtained from the next of kin. This study encompasses 25 postmortem brains obtained between June 2019 and February 2020. The basic demographic included age, biological sex, race (Black, Brown, and White), years of education (range 0-18), as well as presence of smoking, hypertension, and diabetes mellitus. The clinical dementia rating (range 0-3), clinical dementia rating sum of boxes (range 0-18), and neuropsychiatry inventory total score (score 0 or 1) were also recorded. A flowchart of acquisition and analysis has been provided in Supplementary Figure 1.

### Postmortem in situ neuroimaging

Within 24 hours postmortem interval (PMI), postmortem in situ MRI of 25 autopsy cases were acquired using a 32-channel head coil (Nova Medical, USA) and a 7T Siemens Magnetom scanner (Siemens, Germany) at the Medical School of the University of Sao Paulo. Structural imaging included acquisition of T1-weighted (T1w) Magnetization-Prepared 2 Rapid Acquisition Gradient Echoes (MP2RAGE) images at a resolution of 0.75 mm isotropic, T2-weighted (T2w) Sampling Perfection with Application optimized Contrast using different flip angle Evolution (SPACE) images at a resolution of 0.75 mm isotropic, and DWI with detailed sequence parameters provided in Supplementary Table 1. Eight of the cases did not have complete DWI data available.

### Neuroimage scoring

After postmortem imaging, Fazekas score (range 0-3) was utilized to classify the size and confluence of white matter lesions in the periventricular and deep white matter ^34^. MTA score (range 0-4), also known as Scheltens’ scale, was used to classify the width of the choroid fissure and the temporal horn of the lateral ventricle as well as the height of the hippocampus ^35, 36^. Entorhinal cortical atrophy (ERICA) score (range 0-3) has been developed in 2018 as an alternative to the MTA score to identify patients with AD by evaluating the volume loss in the entorhinal cortex ^37^. Koedam score (range 0-3) was to assess parietal atrophy viewed in axial, sagittal, and coronal planes ^38, 39^. The presence of lacunar infarcts less than 15 mm in diameter in the distal distribution of deep penetrating vessels was also recorded ^40^. Basilar artery wall thickness (0.2-0.3 mm in healthy controls ^41^ and 1.2 mm in stroke patients ^42^) and basilar artery diameter (3.2-4.3 mm in healthy controls ^43^ and 4.5 mm in those with artery dilatation ^44^) were also measured. All scoring was provided by a practicing neuroradiologist with twenty years of experience (M.G.M.).

### Neuroimage segmentation

To segment white matter hyperintensity, MP2RAGE and SPACE images were co-registered and manual segmentation of the WMH was performed using ITK-SNAP (version 4.2.0) ^45^. To ensure accuracy, all manual segmentations by one author (J.L.) have been verified by a practicing neuroradiologist (M.G.M.) before exporting the volumes. To evaluate white matter microstructural changes within the WMH, DWI image was co-registered with T1w image using rigid registration in the ITK-SNAP package. The MD was derived from the apparent diffusion coefficient (ADC) maps generated by the scanner and distortion-corrected using the TOPUP tool ^46, 47^. The mean and standard deviation of MD within the ventricles and eyes were obtained from manually labeled regions of interest for normalization of MD within WMH, to account for temperature differences as shown in Supplementary Figure 2. To obtain hippocampal, amygdala, and intracranial volumes, MP2RAGE images were segmented automatically using FreeSurfer (version 8.0). One case did not have complete MP2RAGE slices for FreeSurfer segmentation.

### Neuropathological diagnosis

Tissue sampling and staining included all brain regions recommended by the 2012 National Institutes of Aging - Alzheimer’s Association (NIA-AA) consensus criteria for the neuropathological evaluation of Alzheimer’s disease ^48, 49^.

Immunohistochemical staining for beta amyloid was performed to determine Thal phase ^50^. Phospho-Tau staining was performed to determine Braak NFT stage ^51^. Modified Bielschowsky stains were used to assess neuritic plaque density by Consortium to Establish a Registry for Alzheimer’s disease (CERAD) criteria ^52^. Neuritic plaque scores were based on p-Tau stains. All cases were assigned ABC scores and the subsequent Alzheimer’s disease neuropathologic change (ADNC) rating following the NIA-AA criteria, with details of antibodies provided in Supplementary Table 2. PART was characterized by neurofibrillary tangles in the medial temporal lobe, basal forebrain, brainstem, and olfactory areas including bulb and cortex, in the absence of beta-amyloid plaques ^53^. The presence of aging-related tau astrogliopathy (ARTAG) was based on the existing criteria ^54, 55^. ARTAG was recorded by region and type per consensus recommendations; however, pattern-level distributions (for example, subpial-only vs perivascular) were not uniformly tabulated across the cohort. TDP_-_43 immunohistochemistry was carried out on sections from the amygdala, hippocampus_-_mesial temporal cortex, and midfrontal neocortical regions following consensus guidelines ^56^. TDP-43-positive cases were given a diagnosis of limbic-predominant age-related TDP-43 encephalopathy neuropathological change (LATE-NC). LATE-NC stage was determined following published guidelines by classifying cases based on brain region involvement into stage 1 (amygdala only), stage 2 (stage 1LJ+LJhippocampus and/or entorhinal/transentorhinal cortex), and stage 3 (stage 2LJ+LJmidfrontal cortex). Hippocampal sclerosis (HS) was evaluated unilaterally in the left hemisphere at two coronal sections, one from the anterior hippocampus and the other from the mid_-_hippocampus at the level of the lateral geniculate body. Presence of HS was determined based on severe neuronal loss and gliosis in CA1 and/or subiculum, disproportionate to ADNC in the same regions ^48, 57^. Lewy-type pathology was graded based on immunostaining of alpha synuclein according to the Braak staging system for Parkinson disease and classified as Lewy body disease when Braak stage for Parkinson disease 3 or higher ^58^. Assessment of CAA was based on the beta-Amyloid immunohistochemical signal ^59^: NA, not applicable; no, absent; type 1, signal in cortical capillaries, leptomeningeal and cortical arteries, arterioles, veins, and venules; and type 2, signal in leptomeningeal and cortical vessels, with the exception of cortical capillaries. All scoring was provided by a practicing neuropathologist with twenty years of experience (R.D.R.).

### Statistical analysis

All statistical analyses were conducted using Prism (version 10.2.0.392) unless specified otherwise. Unpaired t-tests were utilized to compare presence and absence of a pathology. Simple linear regression was performed to assess the goodness of fit and the resulting 95% confidence interval, R-squared, p-value have been included in the graph.

Stepwise regression was conducted using SPSS (version 31.0) to explore which independent variables contribute the most to postmortem volumes. The dependent variable was the WMH volume, or hippocampal volume, or amygdala volume; the five independent variables included age, years of education, Thal phase, Braak NFT stage, and CERAD score. The resulting R square, unstandardized coefficients, and associated p-values were reported.

## Results

Within our cohort of 25 people, the average age was 73.0±13.9 (range 53-97) years, with 56% male and 44% female; 72% White, 16% Brown, and 12% Black. Most received 7.4±6.1 (range 0-18) years of formal education when 12, 16, and 18 years of education are equivalent to a high school diploma, a bachelor’s degree, and a master’s degree, respectively. Of relevance to cerebrovascular health, 40% had history of smoking, 24% had hypertension, 32% had diabetes mellitus. In terms of cognition, 72% were cognitively normal (CDR=0), 12% had mild dementia (CDR=1), 4% had moderate dementia (CDR=2), and 12% had severe dementia (CDR=3).

Postmortem neuroimaging assessments performed by a board-certified neuroradiologist were summarized **(Table 1)**. The mean PMI was 15.1±4.3 (range 10-23) hours. Within the cohort, 40% of cases were rated as Fazekas score 3, 16% as Fazekas score 2, 32% as Fazekas score 1, and 12% as Fazekas score 0, indicating that the majority exhibited mild-to-severe WMH at the time of autopsy. Regarding medial temporal lobe atrophy, 36% of participants received an MTA score 0, 24% an MTA score 1, and 40% an MTA score of 2 or higher. ERICA scores were predominantly concentrated at score 0 (40%) and score 1 (36%), while 24% exhibited scores of 2 or 3. The Koedam scale, reflecting parietal atrophy, showed 28% with a score of 0, 44% with a score of 1, and 28% with a score of 2. Lacunar infarcts were present in 44% of the cases.

**Table 1.** Neuroimaging scores by one experienced neuroradiologist.

|  | Fazekas | MTA | ERICA | Koedam<br>(parietal<br>atrophy score) | Lacuna (0 no,<br>1 yes) | Microhemorrhages<br>(0 no, 1 yes) | Basilar artery<br>wall thickness<br>(mm) | Basilar artery<br>diameter (mm) |
| --- | --- | --- | --- | --- | --- | --- | --- | --- |
| #01 | 1 | 0 | 0 | 0 | 0 | 0 | 0.5 | na |
| #02 | 2 | 2 | 1 | 1 | 0 | 1 | 0.5 | na |
| #03 | 3 | 4 | 3 | 2 | 0 | 0 | na | na |
| #04 | 3 | 2 | 1 | 2 | 1 | 0 | 0.6 | 5.2 |
| #05 | 3 | 1 | 1 | 1 | 0 | 0 | 0.8 | 4.7 |
| #06 | 3 | 3 | 3 | 1 | 1 | 1 | 0.5 | 4.2 |
| #07 | 1 | 1 | 1 | 2 | 1 | 0 | 0.4 | 3.2 |
| #08 | 1 | 0 | 0 | 0 | 1 | 0 | 0.4 | 4 |
| #09 | 3 | 4 | 3 | 2 | 1 | 1 | 0.6 | 3.6 |
| #10 | 3 | 2 | 1 | 1 | 1 | 0 | na | 4 |
| #11 | 1 | 0 | 0 | 1 | 0 | 0 | 0.5 | 3.5 |
| #12 | 1 | 1 | 1 | 1 | 0 | 0 | 0.5 | 3.2 |
| #13 | 3 | 2 | 2 | 2 | 1 | 0 | 0.7 | 4.1 |
| #14 | 0 | 0 | 0 | 1 | 0 | 0 | na | 2.3 |
| #15 | 2 | 2 | 1 | 2 | 1 | 0 | 0.5 | 3.5 |
| #16 | 2 | 0 | 0 | 0 | 0 | 0 | na | 2.7 |
| #17 | 3 | 1 | 1 | 1 | 1 | 0 | 0.7 | 4.5 |
| #18 | 0 | 0 | 0 | 0 | 0 | 0 | 0.4 | 3.8 |
| #19 | 3 | 3 | 2 | 2 | 0 | 0 | na | 3.2 |
| #20 | 0 | 1 | 0 | 1 | 1 | 0 | 0.4 | 4 |
| #21 | 1 | 0 | 0 | 0 | 0 | 0 | na | 3.6 |
| #22 | 2 | 3 | 2 | 1 | 0 | 1 | 0.5 | 3.6 |
| #23 | 1 | 0 | 0 | 0 | 0 | 0 | na | 3.3 |
| #24 | 3 | 1 | 1 | 1 | 1 | 0 | na | 2.5 |
| #25 | 1 | 0 | 0 | 0 | 0 | 0 | na | 2.4 |

Abbreviations: ERICA, entorhinal cortical atrophy; MTA, medial temporal atrophy; na, not available.

Vessel measurements demonstrated a mean wall thickness of 0.53±0.12mm. Presence of diabetes (p=0.72), hypertension (p=0.10), or smoking (p=0.96) was not associated with basilar artery thickness. The mean wall diameter was 3.60±0.74mm. Presence of diabetes (p=0.52), hypertension (p=0.91), or smoking (p=0.97) was not associated with basilar artery wall diameter.

The total WMH volume, MD within WMH, ventricles, and eyes, as well as the bilateral hippocampal and amygdala volumes and intracranial volume were summarized **(Table 2)**. The total WMH volume was 14989±15288 (range 98-45480) mm3. The average MD was 0.2±1.0, 1.9±0.1, and 1.7±0.1 x10-3 mm2/s at WMH, ventricles, and eyes, respectively. The resulting ratio of MD at WMH to MD at ventricles was 0.1040±0.0242 whereas ratio of MD at WMH to MD at eyes was 0.1614±0.2169, suggesting normalization to the value at ventricles was less variable. The left and right amygdala volume was 1146.5±414.8 and 1353.2±351.6 mm3, respectively. The left and right hippocampal volume was 3247.6±622.7 and 3337.2±704.4 mm3, respectively. No significant difference between left and right was detected. The estimated intracranial volume was 1351.7±231.6 cm3 across the entire cohort.

**Table 2.** Neuroimaging measures quantified from postmortem MRI.

|  | Total WMH (mm3) | MD WMH (mm2/s) | MD ventricles (mm2/s) | MD eyes (mm2/s) | MD ratio ventricles | MD ratio eyes | Right Amygdala (mm3) | Left Amygdala (mm3) | Right Hippocampus (mm3) | Left Hippocampus (mm3) | ICV (cm3) |
| --- | --- | --- | --- | --- | --- | --- | --- | --- | --- | --- | --- |
| #01 | 5578 | na | na | na | na | na | 1226 | 996.3 | 3710.9 | 3656 | 1333.313 |
| #02 | 13370 | 0.0002 | 0.0020 | 0.0020 | 0.1000 | 0.1000 | 1807 | 1612.9 | 4117.2 | 3903.7 | 1623.6529 |
| #03 | 41850 | 0.0002 | 0.0019 | 0.0019 | 0.1053 | 0.1053 | 1957.3 | 2320 | 2608.5 | 3154.4 | 983.902 |
| #04 | 39390 | na | na | na | na | na | 1353.8 | 1072.4 | 3173.7 | 3381.5 | 1521.83978 |
| #05 | 14040 | 0.0002 | 0.0021 | 0.0022 | 0.0952 | 0.0909 | 1266 | 1153.2 | 3584.6 | 3336.3 | 1618.2798 |
| #06 | 45480 | na | na | na | na | na | 644.8 | 654.9 | 1600.3 | 2104.6 | 1512.5558 |
| #07 | 2847 | 0.0002 | 0.0019 | 0.0019 | 0.1053 | 0.1053 | 1396.4 | 1110.2 | 3436.2 | 3273.5 | 1351.6946 |
| #08 | 4736 | 0.0002 | 0.0021 | 0.0015 | 0.0952 | 0.1333 | 1579.2 | 1334.9 | 3550.3 | 3688.4 | 815.7839 |
| #09 | 15630 | 0.0002 | 0.0018 | 0.0018 | 0.1111 | 0.1111 | 1646.9 | 752.6 | 2460.7 | 2118.9 | 1509.1581 |
| #10 | 31840 | 0.0002 | 0.0022 | 0.0022 | 0.0909 | 0.0909 | 927.2 | 886.5 | 2744.6 | 2897.4 | 1143.666 |
| #11 | 1661 | 0.0002 | 0.0016 | 0.0018 | 0.1250 | 0.1111 | 1555.8 | 1350.8 | 3444.3 | 3260.9 | 1286.2461 |
| #12 | 2510 | 0.0002 | 0.0017 | 0.0019 | 0.1176 | 0.1053 | 1433.3 | 1231.1 | 3908 | 3843 | 1497.407 |
| #13 | 33080 | 0.0002 | 0.0018 | 0.0016 | 0.1111 | 0.1250 | 1215.8 | 742.6 | 3154.3 | 2467.9 | 1432.2222 |
| #14 | 1460 | 0.0002 | 0.0020 | 0.0021 | 0.1000 | 0.0952 | 1443.4 | 1382.2 | 3714.2 | 3639.1 | 1545.4253 |
| #15 | 8654 | 0.0003 | 0.0020 | 0.0020 | 0.1500 | 0.1500 | 845.3 | 360.5 | 2578.9 | 1963.3 | 925.34864 |
| #16 | 11990 | 0.0001 | 0.0017 | 0.0001 | 0.0588 | 1.0000 | 1362.9 | 1255.9 | 3701.1 | 3570.1 | 1251.1572 |
| #17 | 24910 | na | na | na | na | na | 1348.2 | 1331.1 | 3434.7 | 3279.1 | 1152.2341 |
| #18 | 287.3 | na | na | na | na | na | 1598.2 | 1194.3 | 3500.1 | 3663.3 | 1552.897 |
| #19 | 28220 | na | na | na | na | na | na | na | na | na | na |
| #20 | 1666 | 0.0002 | 0.0016 | 0.0017 | 0.1250 | 0.1176 | 1322.8 | 1153.6 | 3834.3 | 3643.4 | 1460.8744 |
| #21 | 1449 | na | na | na | na | na | 1775.8 | 1465.7 | 4539.2 | 4139.1 | 1611.7088 |
| #22 | 5407 | 0.0002 | 0.0022 | 0.0017 | 0.0909 | 0.1176 | 583.9 | 384.6 | 1877 | 2292.9 | 1167.3187 |
| #23 | 1396 | 0.0002 | 0.0015 | 0.0017 | 0.1333 | 0.1176 | 1768.3 | 1602.5 | 4079.9 | 3918.6 | 1614.0705 |
| #24 | 37170 | na | na | na | na | na | 1054.1 | 1058.6 | 3688.6 | 3345.6 | 1222.6226 |
| #25 | 97.67 | 0.0001 | 0.0019 | 0.0015 | 0.0526 | 0.0667 | 1365.4 | 1109 | 3652.2 | 3400.7 | 1306.472 |

Abbreviations: ICV, intracranial volume; MD, mean diffusivity; na, not available; WMH, white matter hyperintensity.

The neuropathological measures were summarized **(Table 3)**. The fresh brain weight was 1230.7±172.9 (range 936-1498) grams in our cohort. Thal phase ranged between 0 and 5, Braak NFT stage ranged between 0 and 6, CERAD score was between 0 and 3 in our cohort. LATE-NC was present in two brains (case #03 and #19), both stage 2 indicating TDP-43 pathology in amygdala and hippocampus. HS was only detected in one postmortem brain (case #03) where the participant also had Thal phase 2, Braak NFT stage 3, and CERAD score 3.

**Table 3.**
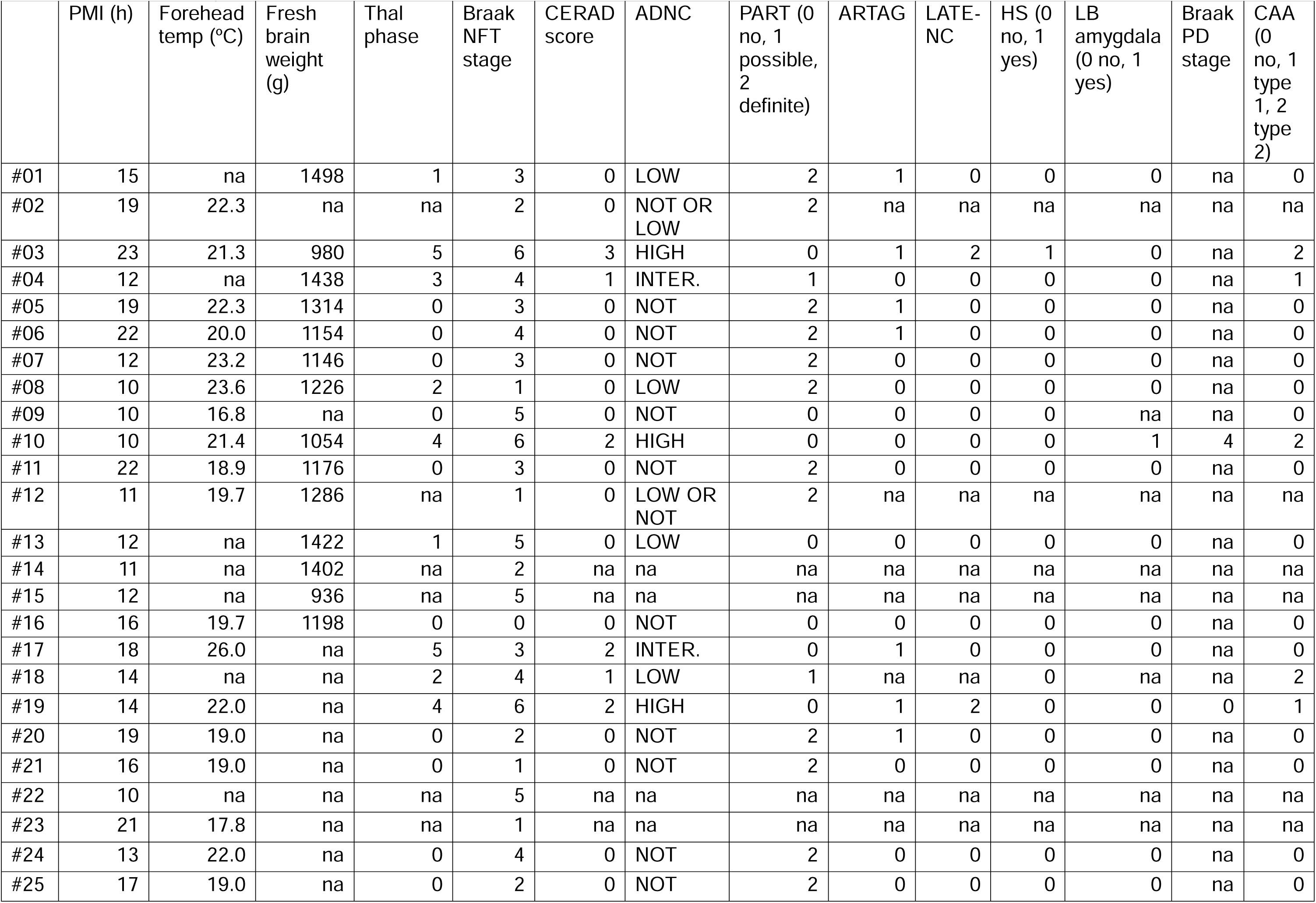
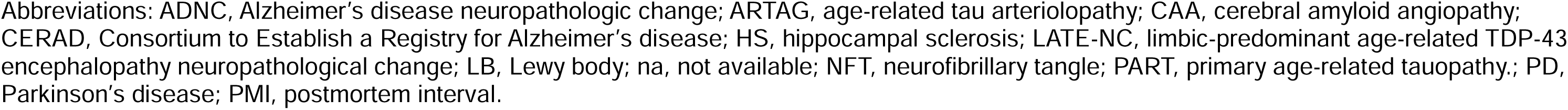
Neuropathological measures of 25 subjects. Arteriolosclerosis and atherosclerosis information was not available.

|  | PMI (h) | Forehead temp (°C) | Fresh brain weight (g) | Thal phase | Braak NFT stage | CERAD score | ADNC | PART (0 no, 1 possible, 2 definite) | ARTAG | LATE-NC | HS (0 no, 1 yes) | LB amygdala (0 no, 1 yes) | Braak PD stage | CAA (0 no, 1 type 1, 2 type 2) |
| --- | --- | --- | --- | --- | --- | --- | --- | --- | --- | --- | --- | --- | --- | --- |
| #01 | 15 | na | 1498 | 1 | 3 | 0 | LOW | 2 | 1 | 0 | 0 | 0 | na | 0 |
| #02 | 19 | 22.3 | na | na | 2 | 0 | NOT OR LOW | 2 | na | na | na | na | na | na |
| #03 | 23 | 21.3 | 980 | 5 | 6 | 3 | HIGH | 0 | 1 | 2 | 1 | 0 | na | 2 |
| #04 | 12 | na | 1438 | 3 | 4 | 1 | INTER. | 1 | 0 | 0 | 0 | 0 | na | 1 |
| #05 | 19 | 22.3 | 1314 | 0 | 3 | 0 | NOT | 2 | 1 | 0 | 0 | 0 | na | 0 |
| #06 | 22 | 20.0 | 1154 | 0 | 4 | 0 | NOT | 2 | 1 | 0 | 0 | 0 | na | 0 |
| #07 | 12 | 23.2 | 1146 | 0 | 3 | 0 | NOT | 2 | 0 | 0 | 0 | 0 | na | 0 |
| #08 | 10 | 23.6 | 1226 | 2 | 1 | 0 | LOW | 2 | 0 | 0 | 0 | 0 | na | 0 |
| #09 | 10 | 16.8 | na | 0 | 5 | 0 | NOT | 0 | 0 | 0 | 0 | na | na | 0 |
| #10 | 10 | 21.4 | 1054 | 4 | 6 | 2 | HIGH | 0 | 0 | 0 | 0 | 1 | 4 | 2 |
| #11 | 22 | 18.9 | 1176 | 0 | 3 | 0 | NOT | 2 | 0 | 0 | 0 | 0 | na | 0 |
| #12 | 11 | 19.7 | 1286 | na | 1 | 0 | LOW OR NOT | 2 | na | na | na | na | na | na |
| #13 | 12 | na | 1422 | 1 | 5 | 0 | LOW | 0 | 0 | 0 | 0 | 0 | na | 0 |
| #14 | 11 | na | 1402 | na | 2 | na | na | na | na | na | na | na | na | na |
| #15 | 12 | na | 936 | na | 5 | na | na | na | na | na | na | na | na | na |
| #16 | 16 | 19.7 | 1198 | 0 | 0 | 0 | NOT | 0 | 0 | 0 | 0 | 0 | na | 0 |
| #17 | 18 | 26.0 | na | 5 | 3 | 2 | INTER. | 0 | 1 | 0 | 0 | 0 | na | 0 |
| #18 | 14 | na | na | 2 | 4 | 1 | LOW | 1 | na | na | 0 | na | na | 2 |
| #19 | 14 | 22.0 | na | 4 | 6 | 2 | HIGH | 0 | 1 | 2 | 0 | 0 | 0 | 1 |
| #20 | 19 | 19.0 | na | 0 | 2 | 0 | NOT | 2 | 1 | 0 | 0 | 0 | na | 0 |
| #21 | 16 | 19.0 | na | 0 | 1 | 0 | NOT | 2 | 0 | 0 | 0 | 0 | na | 0 |
| #22 | 10 | na | na | na | 5 | na | na | na | na | na | na | na | na | na |
| #23 | 21 | 17.8 | na | na | 1 | na | na | na | na | na | na | na | na | na |
| #24 | 13 | 22.0 | na | 0 | 4 | 0 | NOT | 2 | 0 | 0 | 0 | 0 | na | 0 |
| #25 | 17 | 19.0 | na | 0 | 2 | 0 | NOT | 2 | 0 | 0 | 0 | 0 | na | 0 |

Abbreviations: ADNC, Alzheimer's disease neuropathologic change; ARTAG, age-related tau arteriopathy; CAA, cerebral amyloid angiopathy; CERAD, Consortium to Establish a Registry for Alzheimer's disease; HS, hippocampal sclerosis; LATE-NC, limbic-predominant age-related TDP-43 encephalopathy neuropathological change; LB, Lewy body; na, not available; NFT, neurofibrillary tangle; PART, primary age-related tauopathy.; PD, Parkinson's disease; PMI, postmortem interval.

Lewy body pathology was detected in case #10, amygdala-predominant Lewy body pathology and Braak PD stage 4. For CAA, 14 cases had no CAA, two cases were CAA type 1, and three other cases were CAA type 2. Data for arteriolosclerosis and atherosclerosis was not available.

Higher postmortem Fazekas scores **(Figure 1)** correlate with older age (R^2^=0.4375, p=0.0003), fewer years of education (R^2^=0.2571, p=0.0097), presence of hypertension (p=0.0325), higher basilar artery wall thickness (R^2^=0.6426, p=0.0002), and higher Braak NFT stage (R^2^=0.3571, p=0.0016). No significant differences were observed for PMI (p=0.95), presence of diabetes (p=0.79) or smoking (p=0.22), presence of lacuna (p=0.08), Thal phase (p=0.10), CERAD score (p=0.06), presence of PART (p=0.22), ARTAG (p=0.22), or CAA (p=0.21).

**Figure 1.**
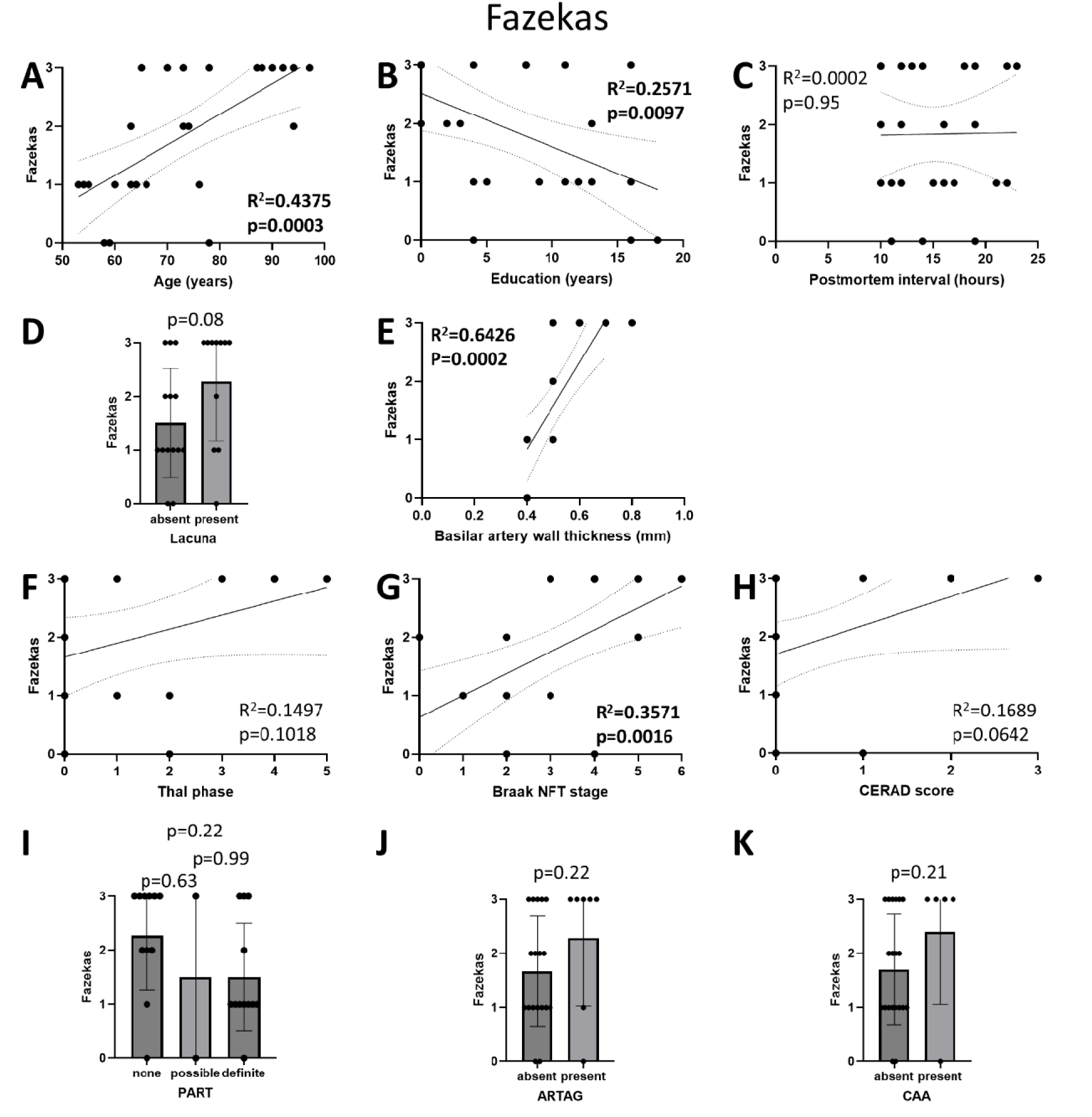
Higher postmortem Fazekas scores correlate with (A) older age, (B) fewer years of education, (E) higher basilar artery wall thickness, and (G) higher Braak NFT stage. No significant differences are observed for (C) PMI, (D) presence of lacuna, (F) Thal phase, (H) CERAD score, presence of (I) PART, (J) ARTAG, or (K) CAA.

Similar to Fazekas score, higher postmortem WMH volumes **(Figure 2)** correlated with older age (R^2^=0.3466, p=0.0020), fewer years of education (R^2^=0.2123, p=0.0205), presence of hypertension (p=0.0149), higher Fazekas score (R^2^=0.7205, p<0.0001), higher basilar artery wall thickness (R^2^=0.2720, p=0.0383), and higher Braak NFT stage (R^2^=0.3654, p=0.0014). In addition, higher postmortem WMH volumes also correlated with presence of lacuna (p=0.0305), higher Thal phase (R^2^=0.2173, p=0.0443), and higher CERAD score (R^2^=0.2522, p=0.0203), associations that were not detected using the Fazekas score. No significant differences were observed for PMI (p=0.60), presence of diabetes (p=0.54) or smoking (p=0.08), presence of PART (p=0.09), ARTAG (p=0.40) or CAA (p=0.10).

**Figure 2.**
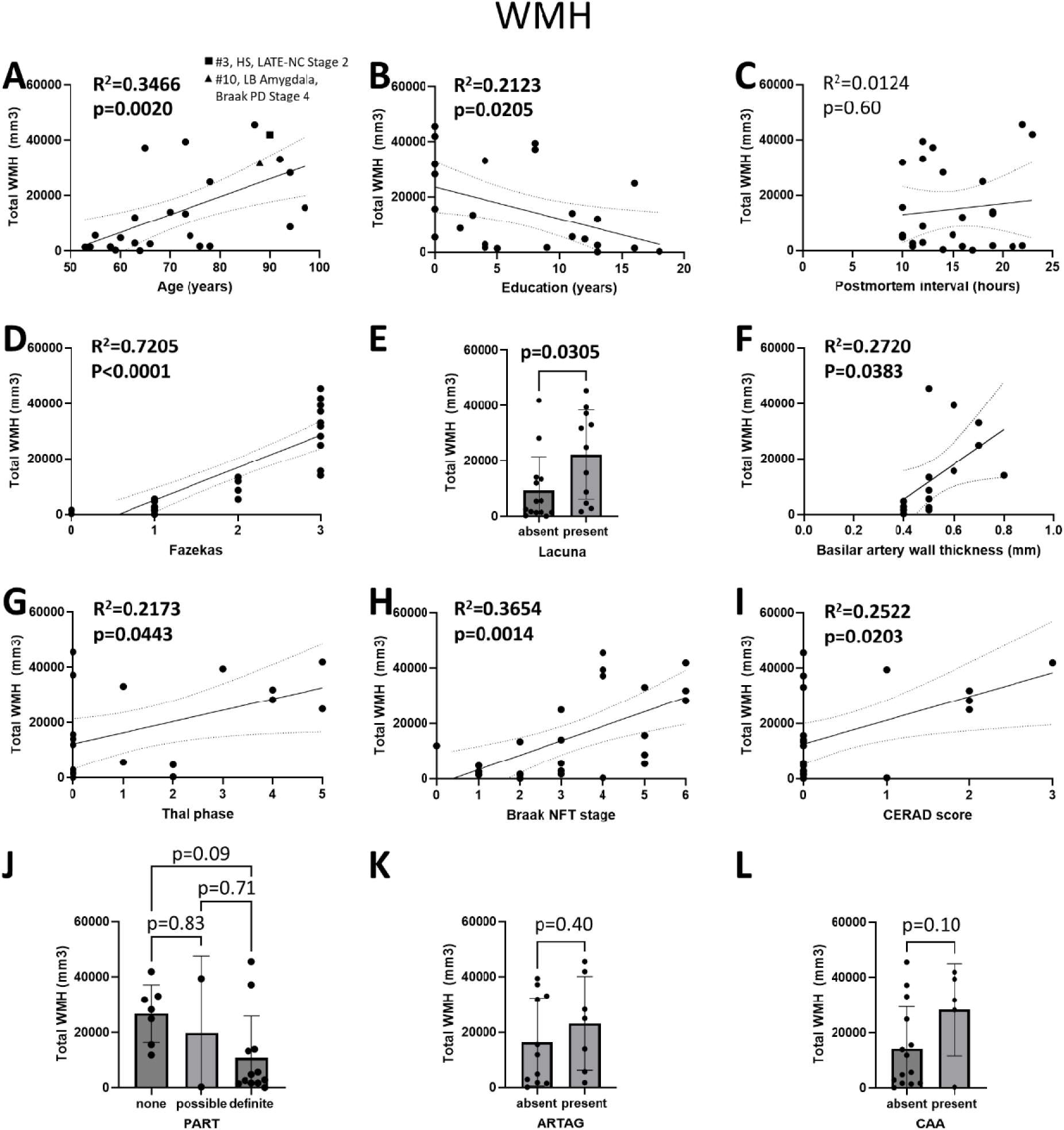
Higher postmortem WMH volumes correlate with (A) older age, (B) fewer years of education, (D) higher Fazekas score, (E) presence of lacuna, (F) higher basilar artery wall thickness, (G) higher Thal phase, (H) higher Braak NFT stage, and (I) higher CERAD score. No significant differences are observed for (C) PMI, (J) PART, (K) ARTAG, or (L) CAA. In our cohort of 25, only case #03 (square) exhibits hippocampal sclerosis and LATE-NC stage 2. Only case #10 (triangle) shows amygdala-predominant Lewy body pathology and Braak PD stage 4.

MD within WMHs **(Figure 3)**, normalized to ventricular MD, did not show significant correlations with age (p=0.22). When normalized to MD within eyes, MD of WMH did not correlate with age either (p=0.49) and had a higher variability compared to ventricular MD. MD of WMH did not correlate with PMI (p=0.75), forehead temperature (p=0.49), Fazekas score (p=0.67), total WMH volume (p=0.81), left hippocampal (p=0.43), right hippocampal (p=0.81), left amygdala (p=0.74), or right amygdala (p=0.85) volumes.

**Figure 3.**
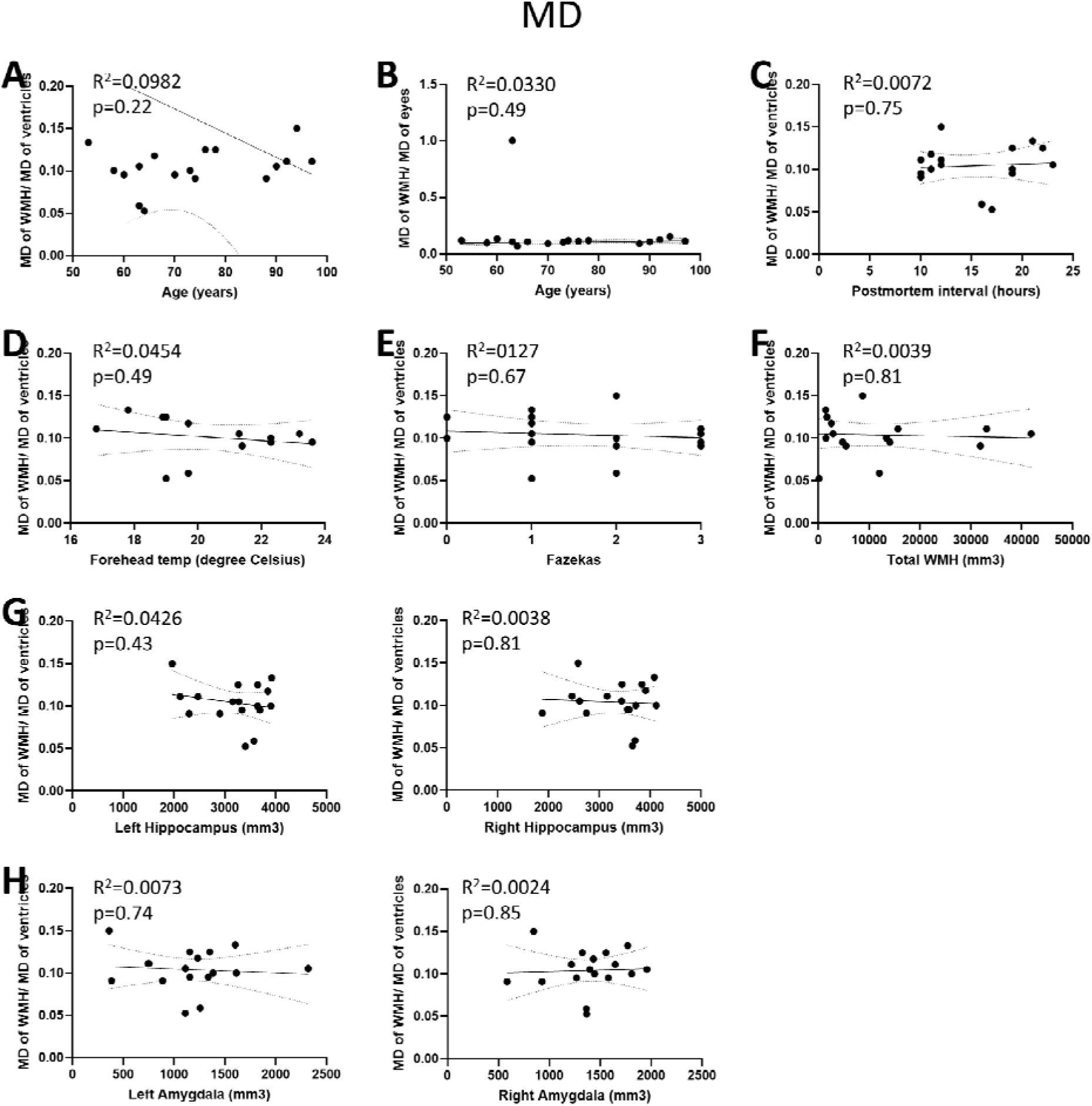
Postmortem MD of WMH normalized by MD of ventricles does not correlate with (A) age, (C) postmortem interval, (D) forehead temperature, (E) Fazekas score, (F) total WMH volume, (G) bilateral hippocampal volumes, or (H) bilateral amygdala volumes. Please note that (B) MD of WMH normalized by MD of eyes shows higher variability compared to normalization of ventricles, does not correlate with any measurements either.

Higher bilateral hippocampal volumes **(Figure 4)** correlated with younger age (Left: R^2^=0.6463, p<0.0001; Right: R^2^=0.4962, p=0.0001), more years of education (Left: R^2^=0.4268, p=0.0005; Right: R^2^=0.4285, p=0.0005), lower MTA score (Left: R^2^=0.5815, p<0.0001; Right: R^2^=0.5069, p<0.0001), lower ERICA score (Left: R^2^=0.5254, p<0.0001; Right: R^2^=0.5912, p<0.0001), lower Koedam score (Left: R^2^=0.3576, p=0.0020; Right: R^2^=0.2421, p=0.0146), and lower Braak NFT stage (Left: R^2^=0.5381, p<0.0001; Right: R^2^=0.5486, p<0.0001). No significant differences were observed for Thal phase (Left: p=0.98; Right: p=0.34), CERAD score (Left: p=0.68; Right: p=0.15), presence of PART (Left: p=0.10; Right: p=0.19), presence of ARTAG (Left: p=0.90; Right: p=0.42), or presence of CAA (Left: p=0.90; Right: p=0.28). No analysis was performed for LATE-NC due to the low prevalence (n=2) in our cohort.

**Figure 4.**
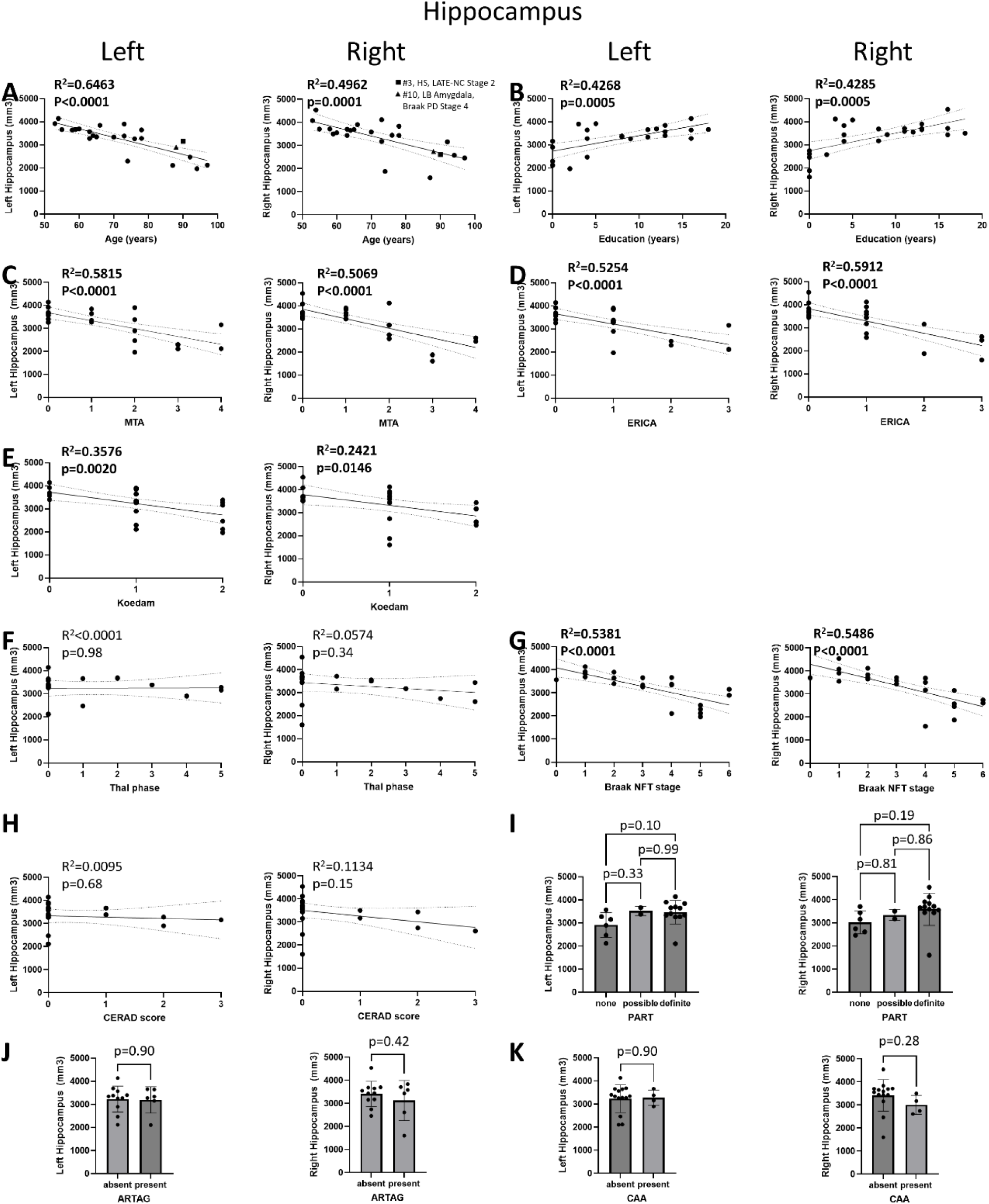
Higher bilateral hippocampal volumes correlate with (A) younger age, (B) more years of education, (C) lower MTA score, (D) lower ERICA score, (E) lower Koedam score, and (G) lower Braak NFT stage. No significant correlations are observed for (F) Thal phase, (H) CERAD score, or presence of (I) PART, (J) ARTAG, or (K) CAA. In our cohort of 25, only case #03 (square) exhibits hippocampal sclerosis and LATE-NC stage 2. Only case #10 (triangle) shows amygdala-predominant Lewy body pathology and Braak PD stage 4.

Higher amygdala volume **(Figure 5)** correlated with higher CERAD score in the left hemisphere (R^2^=0.2380, p=0.0291) but not in the right hemisphere (p=0.49). No significant correlations were observed for age (Left: p=0.11; Right: p=0.15), years of education (Left: p=0.18; Right: p=0.13), MTA score (Left: p=0.32; Right: p=0.29), ERICA score (Left: p=0.34; Right: p=0.25), Koedam score (Left: p=0.34; Right: p=0.49), Thal phase (Left: p=0.06; Right: p=0.48), Braak NFT stage (Left: p=0.10; Right: p=0.06), presence of PART (Left: p=0.99; Right: p=0.97), ARTAG (Left: p=0.41; Right: p=0.58), or CAA (Left: p=0.21; Right: p=0.51).

**Figure 5.**
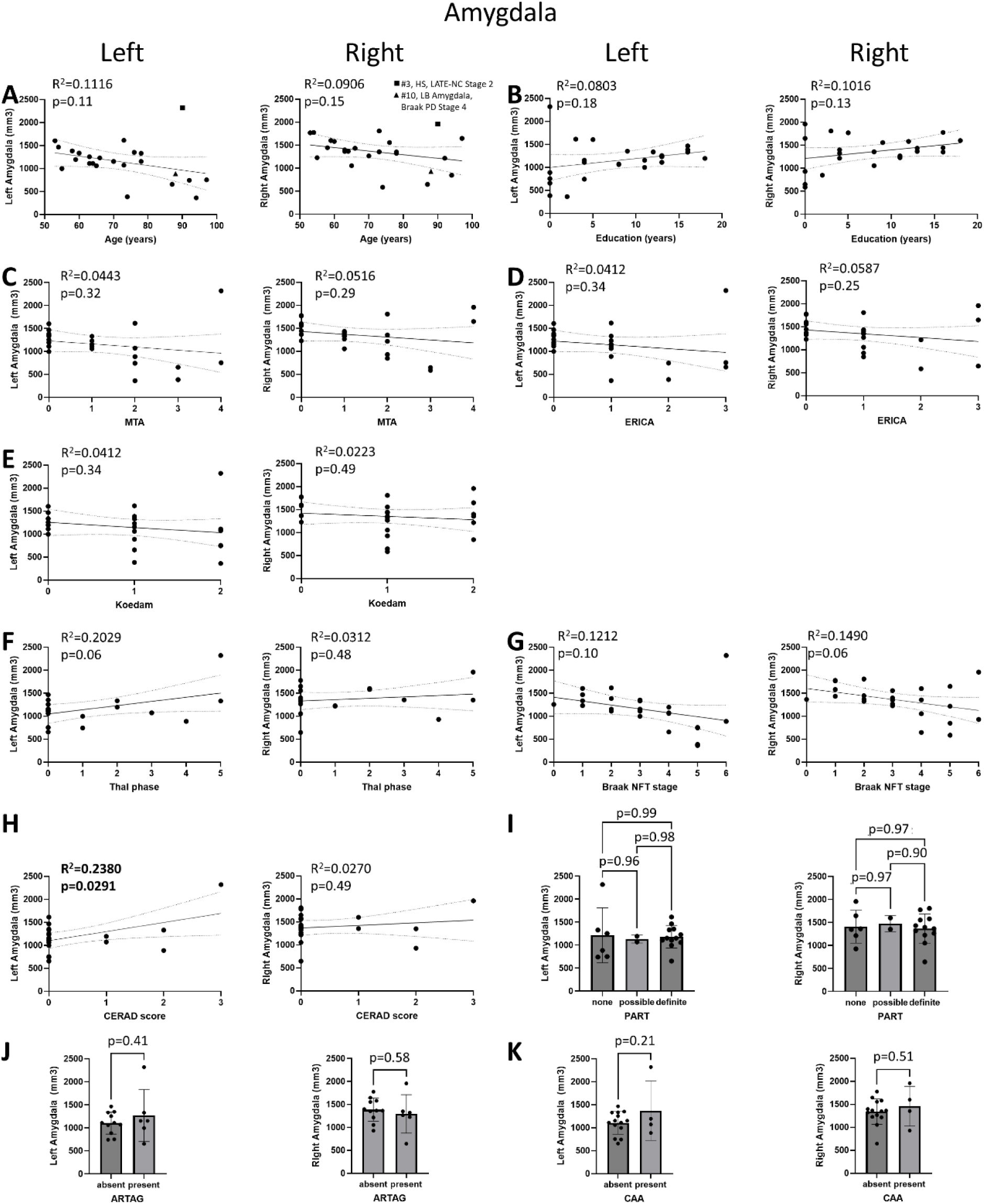
Higher left amygdala volume only correlates with (H) higher CERAD score. No significant correlations are observed for (A) age, (B) education attainment, (C) MTA, (D) ERICA, (E) Koedam, (F) Thal phase, (G) Braak NFT stage, presence of (I) PART, (J) ARTAG, or (K) CAA. In our cohort of 25, only case #03 (square) exhibits hippocampal sclerosis and LATE-NC stage 2. Only case #10 (triangle) shows amygdala-predominant Lewy body pathology and Braak PD stage 4.

Stepwise regression **(Figure 6)** indicated that total WMH volume was primarily driven by Braak NFT stage, accounting for 32.8% of the variance. Left hippocampal volume was associated with age, Thal phase, and Braak NFT stage, with these predictors collectively explaining 85.0% of the variance. In contrast, right hippocampal volume was influenced only by Braak NFT stage, accounting for 52.0% of the variance. Left amygdala volume was predicted by the CERAD score, Braak NFT stage, Thal phase, education, and age. However, no significant model emerged for the right amygdala, suggesting that none of the predictors showed a significant linear relationship with the dependent variable.

**Figure 6.**
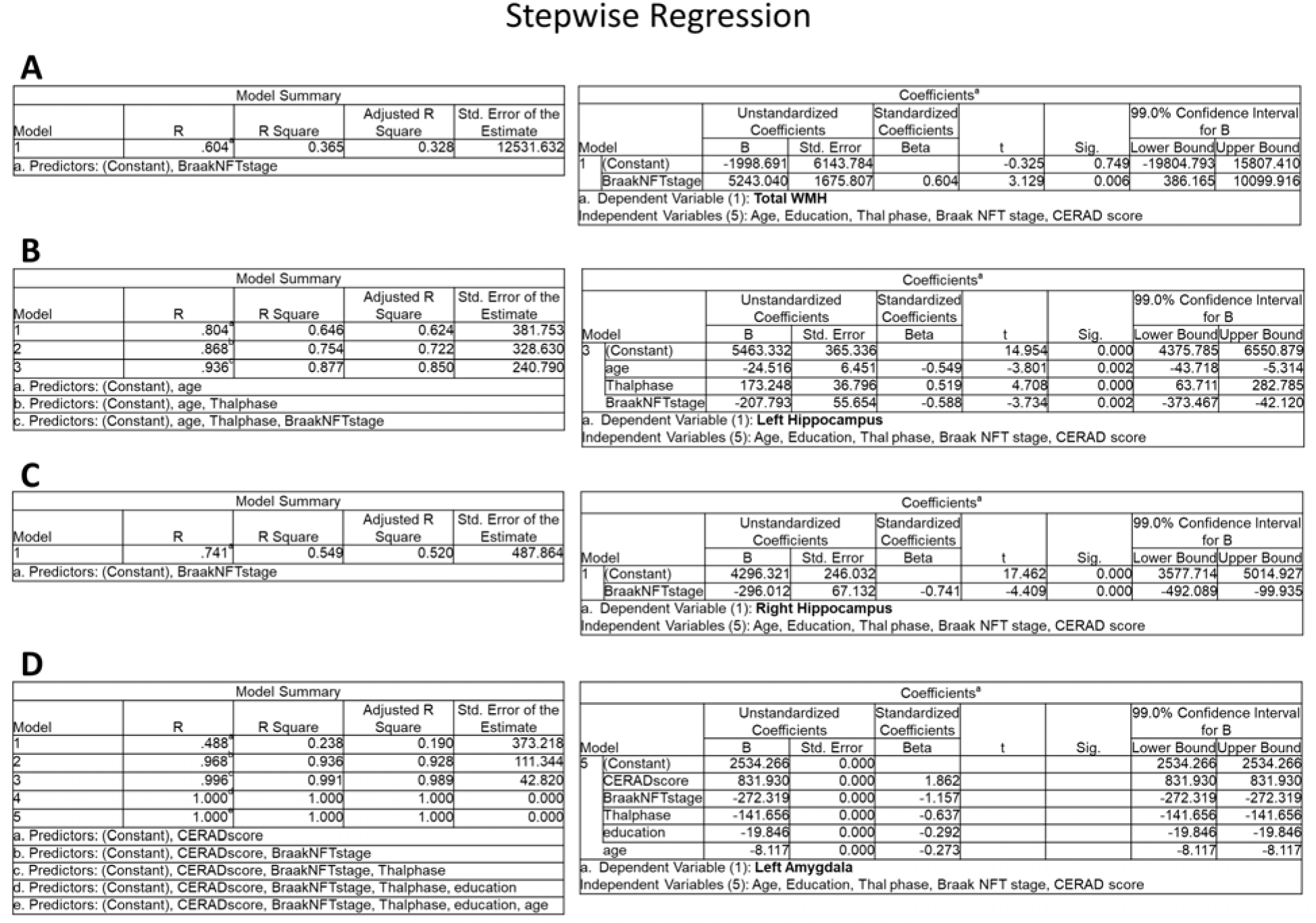
Stepwise regression shows (A) total WMH volume is primarily driven by Braak NFT stage. (B) Left hippocampal volume is driven by age, Thal phase, and Braak NFT stage while (C) right hippocampal volume is only affected by Braak NFT stage. (D) Left amygdala volume is driven by CERAD score, Braak NFT stage, Thal phase, education, and age; however, no significant relationship was found in right amygdala.

## Discussion

In this community cohort of 25 older adults (mean age 73 years), postmortem in situ MRI showed mild-to-severe WMHs and mild-to-moderate medial temporal and global cortical atrophy. Higher WMH volumes were associated with higher Fazekas score, older age, fewer years of education, more advanced AD pathology, whereas WMH mean diffusivity showed no significant associations. In addition, hippocampal volumes were lower in elderly, less educated people and were associated with higher atrophy scores and higher Braak NFT stage, while higher left amygdala volume only correlated with higher CERAD neuritic plaque scores.

In a large memory clinic, in vivo Fazekas score is associated with amyloid PET burden ^60^. However, Fazekas score, after correcting for age and Braak NFT stage, can no longer distinguish PART from AD pathology ^61^, suggesting it may be driven by Braak NFT stage. Here we found that higher postmortem Fazekas scores correlated with older age, fewer years of education, higher basilar artery wall thickness, and higher Braak NFT stage. No significant associations were detected in Thal phase, likely due to the low prevalence of amyloid plaques in our cohort.

Previous studies have shown that greater WMH burden correlate with atrophy in the fronto-insular and medial temporal regions in both AD and PART, with PART demonstrating stronger correlations ^61^ and that higher WMH burden combined with elevated cortical tau levels ^62^, similar to those seen in advanced PART, predicts more severe cognitive symptoms than either factor alone. In our analysis, we did not find any significant relationship between WMH and PART pathology. Two possible explanations exist: (i) regional WMH volumes or spatial WMH patterns ^63^ may reveal associations that are not captured in global WMH volumes and (ii) our cohort is limited by sample size, with 7 non-PART, 2 possible PART, and 12 definite PART cases, leaving the remainder unavailable, which restricts statistical power to detect cross-sectional differences. One study found that postmortem WMH from T2w images is still much more accurate (accuracy 71%) than T1w/T2w ratio (accuracy 42%) and magnetization transfer ratio (accuracy 39%) for assessment of demyelination in the cortical regions ^25^. Whether or not the same applies to the white matter remains to be investigated. In addition to PART pathology, those who had CAA should have a higher WMH volume ^64–66^ and a higher MD ^67^ compared to non-CAA but the WMH spatial pattern ^65^ is similar between CAA, AD/MCI, and healthy aging.

Interestingly, the correlation between PiB PET burden and WMH volume was significant in CAA but not in AD/MCI or healthy aging ^66^. Increased WMH volume in the juxtacortical region is linked to probable CAA diagnosis ^68^. The postmortem WMH volume is significantly correlated with the percentage of cortical CAA area ^69^. In our analysis, we did not see a significant increase in WMH volume in our 5 CAA cases compared to 14 non-CAA cases.

Postmortem in situ imaging as opposed to ex vivo imaging of fixed tissue allows avoidance of tissue shrinkage in the hippocampus, and reduction in MD bias in the white matter^23^ and postmortem MD shows distinction between brains with amyotrophic lateral sclerosis and brains from healthy controls ^24^. In a longitudinal study of 266 participants where they were imaged three times over a course of 9 years, impaired microstructural changes in vivo revealed by an increase in MD preceded conversion into WMH and continuously declined over time, highlighting the potential to visualize more subtle injuries in white matter using MD metrics ^70^. However, in our analysis, MD within WMHs, normalized to ventricular MD did not show significant associations. This suggests that white matter microstructural changes within WMHs are relatively homogeneous across our cohort, and that the intensity of tissue degradation is less relevant than the overall extent of the lesions. Diffusivity is sensitive to temperature, and both forehead and core temperatures have been shown to correlate significantly with MD ^71^. In our cohort, forehead temperature was indeed correlated with ventricular MD (R-squared=0.5308, p=0.0047) but not with WMH MD or with the MD ratio of WMH to ventricle, a finding similar to our previous study in a cohort of 47 cases (ages 31-91) ^72^. Antemortem FA can distinguish postmortem arteriosclerosis between none and mild (and of course between none and moderate as well as between none and severe) ^73^. Postmortem FA in gray matter can differentiate AD brains ^31^ and MS brains ^26^ from control brains.

In AD, significant correlations have been found between hippocampal volume and both Thal phase and CERAD score in postmortem brains ^2^. Here no significant correlations were observed with Thal phase or CERAD score, likely due to most of the cases not being AD cases. Amygdala and hippocampus are the two major regions where LATE-NC begins. In this cohort, only two autopsy cases showed LATE-NC (both stage 2), limiting our ability to assess hippocampal or amygdala volume associations. Interestingly, MTA score has discriminative ability known as area under the curve, comparable to amygdala volume and hippocampal volume, for differentiating LATE-NC from non-LATE-NC cases ^33^.

In our analysis, higher amygdala volume was correlated with higher CERAD scores only in the left hemisphere, but not in the right. No other correlations were observed between amygdala volume and Thal phase or Braak NFT stage, consistent with previous findingsLJ . The presence of TDP-43 pathology in PART has been associated with greater volume loss in the hippocampus and amygdalaLJ while increasing Braak NFT stage is linked to atrophy in anterior and medial temporal regions ^75, 76^. Regarding hemispheric asymmetry, atrophy of the left amygdala was more severe than that in contralateral region in the AD group compared to healthy controls ^77^, consistent with our observations that the left amygdala volume is smaller, though insignificant, than the right amygdala.

This study has three major limitations. First, our sample size is smaller than other postmortem in situ studies ^20, 21^. Data on arteriolosclerosis and atherosclerosis were not available in our cohort. Previous studies have shown that arteriolosclerosis is a primary cause of WMHs ^78^ and one of the major predictors of postmortem amygdala volume in AD and related neurodegenerative pathologies ^2^. Presence of hypertension and diabetes is also linked to both arteriolosclerosis pathology and increased WMH load in the deep frontal region ^68^. Second, other biological and genetic variables, such as ApoE4 status, are unknown and may contribute to the observed WMH and microstructural changes. Third, the MD values derived from ADC maps may not be as sensitive as we expected. One study ^79^ reported that ADC maps are useful for real-time diagnosis of stroke and tumors but the postmortem ADC values can decrease by up to 70% compared to values in healthy living individuals. As PMI increases, ADC values decrease significantly when considering all regions together, particularly in gray matter. We therefore mitigated this issue by normalizing it to ventricular or ocular MD. Also, in situ 7T MRI has a lot of low signals from vessels, making it more difficult to detect microhemorrhage, a biomarker for CAA.

Future efforts will expand on quantification of hippocampal subfield volumes for correlating with neuropathological measures. For instance, postmortem subiculum and entorhinal cortex volumes have been shown to correlate with clinical dementia rating scores, and p-tau immunostaining intensity was identified as the strongest predictor of hippocampal atrophy, particularly in the subiculum ^32^. We will also explore postmortem amygdala nuclei volumes including the lateral nucleus and the basal nucleus ^80^; though current evidence suggests that only the volume of the lateral nucleus can distinguish LATE-NC from non-LATE-NC autopsy cases ^33^. Finally, we will dive into regional WMH volumes and explore WMH spatial patterns.

Postmortem studies have shown that WMH burden correlates with the white matter sclerotic index, a measure of small vessel disease, only in the occipital lobe, but not in the frontal, temporal, or parietal lobes in ADLJ . In FTLD-TDP brains, postmortem WMH volume is higher, particularly in the frontal region, compared to those with AD ^81^.

In summary, higher Fazekas scores were associated with older age, lower education, hypertension, increased basilar artery wall thickness, and higher Braak NFT stage. Increased WMH volume was also correlated with the presence of lacunes, higher Thal phase, and higher CERAD score, associations not observed with Fazekas scores. Hippocampal volumes were lower in older individuals with less education and were associated with higher atrophy scores and greater Braak NFT stage. Larger amygdala volume was associated only with higher CERAD scores. Quantitative MRI may detect neuropathological associations more sensitively than visual ratings. Tau pathology appears to be a key predictor of WMH burden and hippocampal atrophy.

## Data Availability

All data produced in the present study are available upon reasonable request to the corresponding authors.

## Acknowledgements

This study was supported by the National Institute on Aging of the National Institutes of Health under award number R01AG070826, Alzheimer’s Association, and The Foundation of the American Society of Neuroradiology under award number 25IR-AND-1438973. JL is partly supported by the BrightFocus Foundation A2025009F.

## Consent statement

All brain donors consented to participate in this research study.

---

**Supplementary Figure 1.**
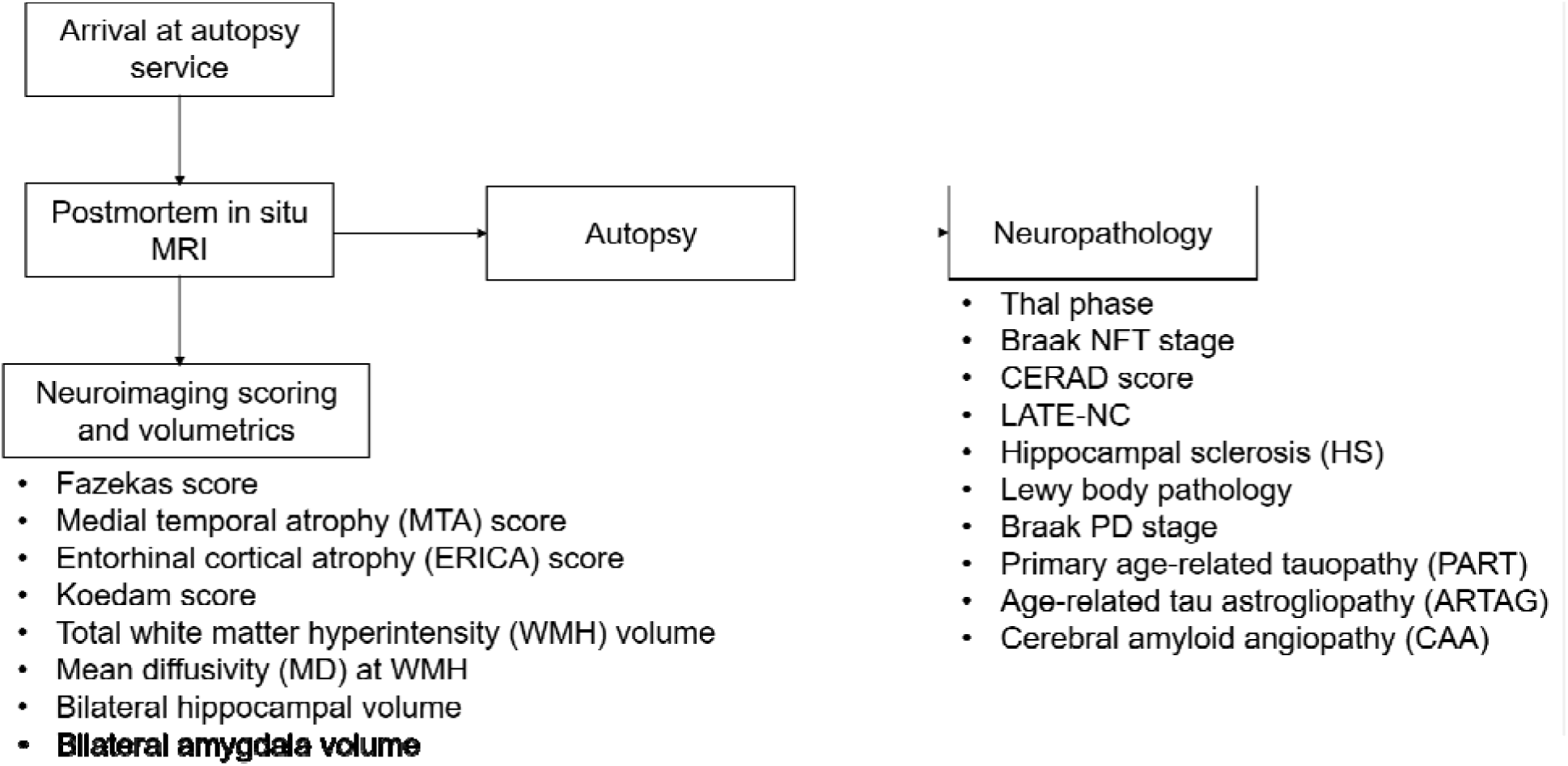
Acquisition and analysis flow chart from postmortem in situ MRI to neuroimaging volumetrics and neuropathological diagnoses.

**Supplementary Figure 2.**
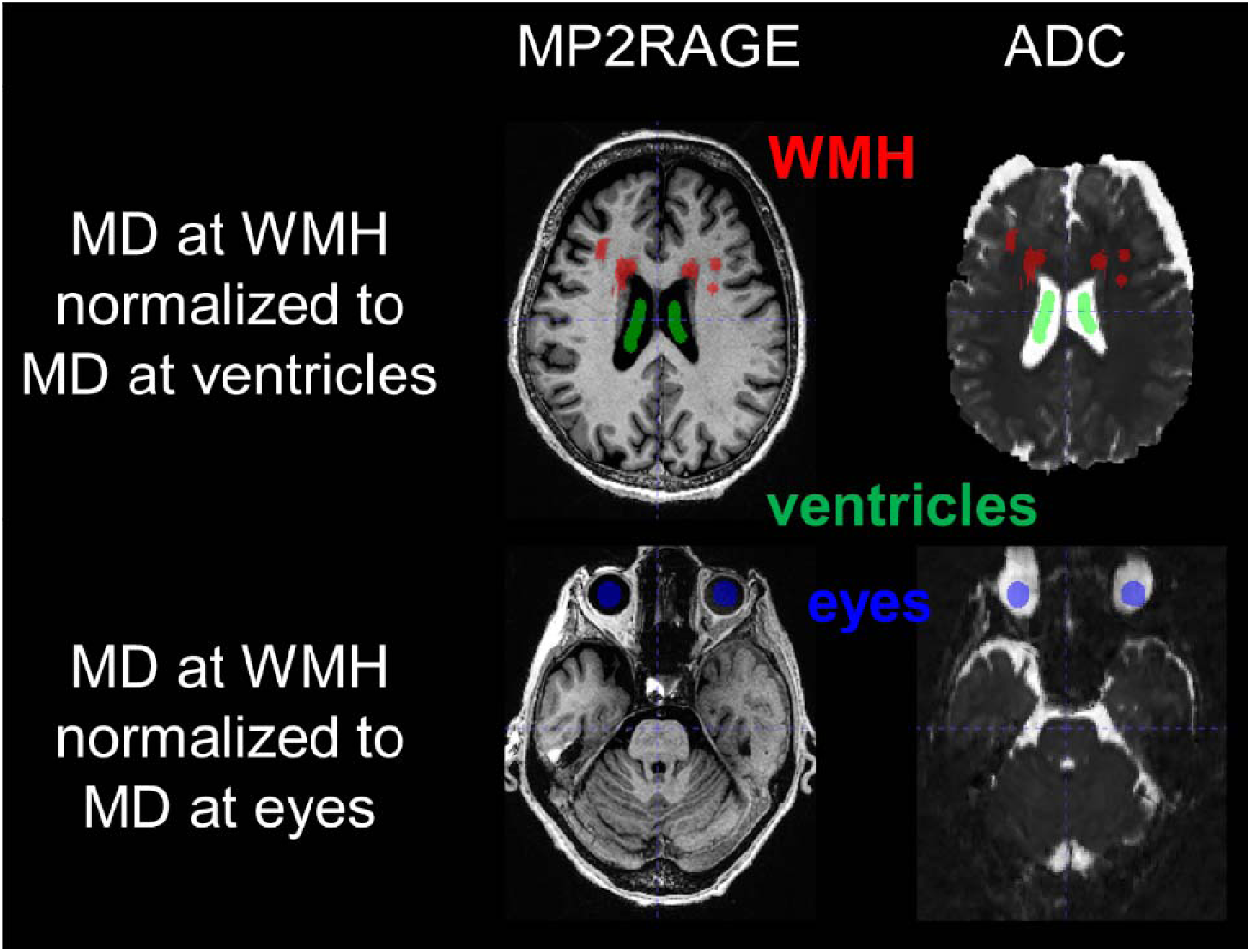
Representative T1-weighted MP2RAGE, Magnetization-Prepared 2 Rapid Acquisition Gradient Echoes (MP2RAGE), apparent diffusion coefficient (ADC), and resliced ADC images for mean diffusivity (MD) at white matter hyperintensity (WMH) normalized to either MD at ventricles or MD at eyes in same subject.

**Supplementary Table 1.** Sequence parameters of 7T postmortem in situ MRI.

|  | T1w MP2RAGE | T2w SPACE | DWI |
| --- | --- | --- | --- |
| Resolution | 0.75 mm iso | 0.75 mm iso | 1.3 mm iso |
| Echo time | 1.93 ms | 91 ms | 74.8 ms |
| Inversion time | 800/2700 ms | n/a | n/a |
| Repetition time | 6000 ms | 2000 ms | 9700 ms |
| Acceleration factor | 3 | 3 | 3 |
| Number of averages | 1 | 1 | 1 |
| Acquisition time | 10 min 51 sec | 14 min 19 sec | 12 min 47 sec |
| Purpose | Intracranial volume, hippocampal volume, amygdala volume | White matter hyperintensity volume | White matter mean diffusivity |

Abbreviations: DWI, diffusion-weighted imaging; MP2RAGE, Magnetization-Prepared 2 Rapid Acquisition Gradient Echoes; n/a, not applicable; SPACE, Sampling Perfection with Application optimized Contrast using different flip angle Evolution; T1w, T1-weighted; T2w, T2-weighted; TSE, Turbo Spin Echo.

**Supplementary Table 2.**
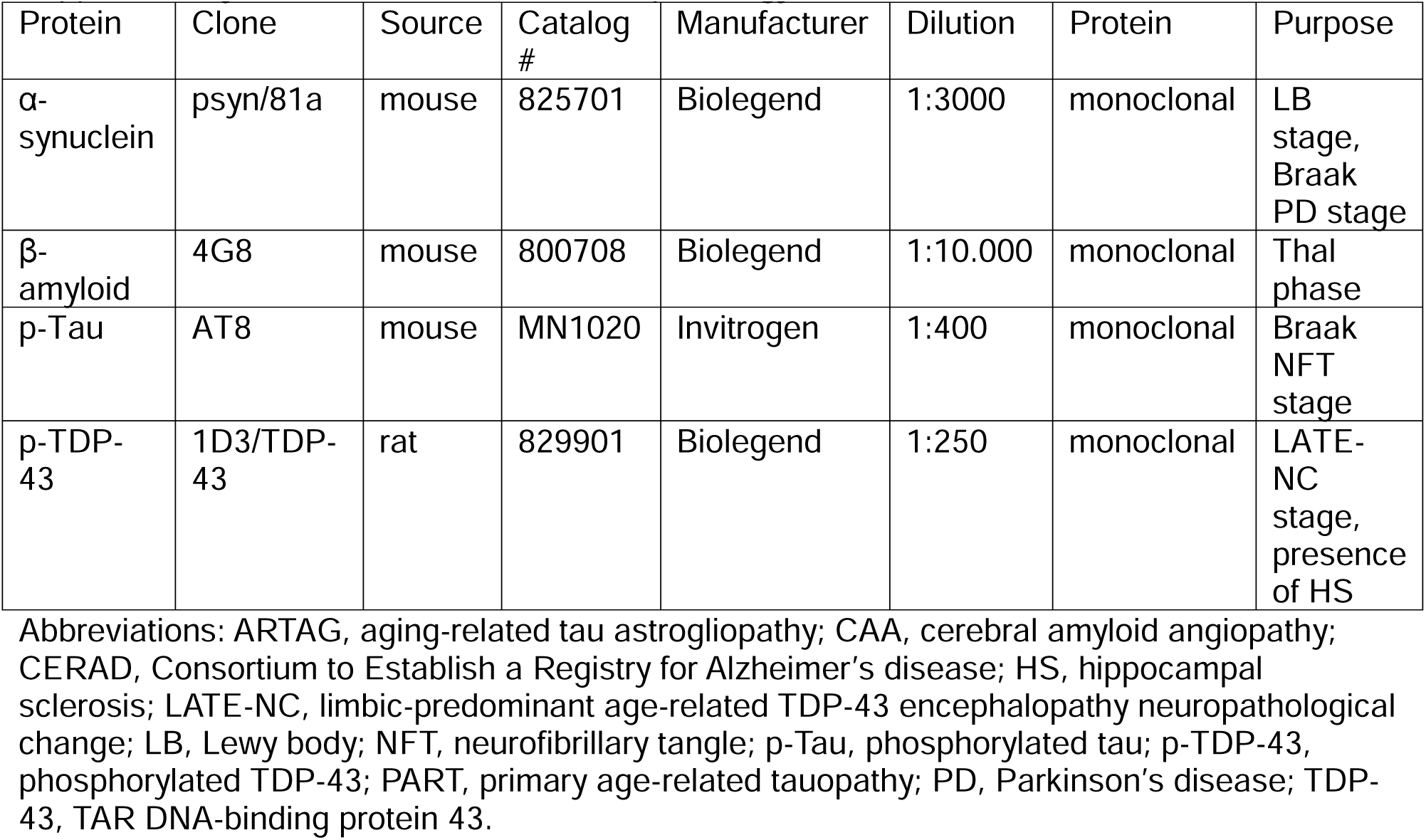
Antibodies for histopathology.

